# Awareness, knowledge and trust in the Greek authorities towards COVID-19 pandemic: results from the Epirus Health Study cohort

**DOI:** 10.1101/2020.11.10.20229146

**Authors:** Afroditi Kanellopoulou, Fotios Koskeridis, Georgios Markozannes, Emmanouil Bouras, Chrysa Soutziou, Konstantinos Chaliasos, Michail T Doumas, Dimitrios E Sigounas, Vasilios T Tzovaras, Agapios Panos, Yiolanda Stergiou, Kassiani Mellou, Dimitrios Papamichail, Eleni Aretouli, Dimitrios Chatzidimitriou, Fani Chatzopoulou, Eleni Bairaktari, Ioanna Tzoulaki, Evangelos Evangelou, Evangelos C Rizos, Evangelia Ntzani, Konstantinos Vakalis, Konstantinos K Tsilidis

**Affiliations:** Department of Hygiene and Epidemiology, University of Ioannina School of Medicine, Ioannina, Greece; Department of Hygiene, Social-Preventive Medicine and Medical Statistics, Aristotle University of Thessaloniki School of Medicine, Thessaloniki, Greece; Ioannina Medical Care, Ioannina, Greece; Directorate of Epidemiological Surveillance and Intervention for Infectious Diseases, Hellenic National Public Health Organization, Athens, Greece; Department of Public Health Policy, School of Public Health, University of West Attica, Athens, Greece; School of the Social Sciences, University of Ioannina, Ioannina, Greece; Laboratory of Cognitive Neuroscience, School of Psychology, Aristotle University of Thessaloniki, Thessaloniki, Greece; Laboratory of Microbiology, Aristotle University of Thessaloniki School of Medicine, Thessaloniki, Greece; Biochemistry Department, University of Ioannina, Ioannina, Greece; Department of Epidemiology and Biostatistics, School of Public Health, Imperial College London, London, UK; Department of Internal Medicine, University Hospital of Ioannina, Ioannina, Greece; School of Medicine, European University of Cyprus, Nicosia, Cyprus; Center for Evidence-Based Medicine, Department of Health Services, Policy and Practice, School of Public Health, Brown University, Providence, RI, USA; Institute of Biosciences, University Research Center of loannina, University of Ioannina, Ioannina, Greece

**Keywords:** COVID-19, Knowledge, Trust in authorities, cohort study, Epirus Health Study, Exposure-wide association analysis

## Abstract

**Background:** To assess the level of knowledge and trust in the policy decisions taken regarding the coronavirus disease (COVID-19) pandemic among Epirus Health Study (EHS) participants.

**Methods:** The EHS is an ongoing and deeply-phenotyped prospective cohort study that has recruited 667 participants in northwest Greece until August 31^st^, 2020. Level of knowledge on coronavirus (SARS-CoV-2) transmission and COVID-19 severity was labeled as poor, moderate or good. Variables assessing knowledge and beliefs towards the pandemic were summarized overall and by gender, age group (25-39, 40-49, 50-59, ≥60 years) and period of report (before the lifting of lockdown measures in Greece: March 30^th^ to May 3^rd^, and two post-lockdown time periods: May 4^th^ to June 31^st^, July 1^st^ to August 31^st^). An exposure-wide association analysis was conducted to evaluate the associations between 153 explanatory variables and participants’ knowledge. Correction for multiple comparisons was applied using a false discovery rate (FDR) threshold of 5%.

**Results:** A total of 563 participants (49 years mean age; 60% women) had available information on the standard EHS questionnaire, the clinical and biochemical measurements, and the COVID-19-related questionnaire. Percentages of poor, moderate and good knowledge status regarding COVID-19 were 4.5%, 10.0% and 85.6%, respectively. The majority of participants showed absolute or moderate trust in the Greek health authorities for the management of the epidemic (90.1%), as well as in the Greek Government (84.7%) and the official national sources of information (87.4%). Trust in the authorities was weaker in younger participants and those who joined the study after the lifting of lockdown measures (p-value≤0.001). None of the factors examined was associated with participants’ level of knowledge after correction for multiple testing.

**Conclusions:** High level of knowledge about the COVID-19 pandemic and trust in the Greek authorities was observed, possibly due to the plethora of good quality publicly available information and the timely management of the pandemic at its early stages in Greece. Information campaigns for the COVID-19 pandemic should be encouraged even after the lifting of lockdown measures to increase public awareness.

## BACKGROUND

In December 2019, a cluster of patients suffering from atypical pneumonia was identified in the city of Wuhan, China (1). The outbreak was caused by a novel coronavirus named Severe Acute Respiratory Syndrome-Coronavirus-2 (SARS-CoV-2), which shares pathogenicity characteristics with Severe Acute Respiratory Syndrome-Coronavirus (SARS-CoV) and Middle East Respiratory Syndrome-Coronavirus (MERS-CoV), both responsible for epidemics caused in the 2010s (1,2). The rapid spread of the virus around the globe led to a huge public health crisis and on March 11^th^, 2020, the World Health Organization (WHO) characterized coronavirus disease (COVID-19) as a pandemic (3). Up to August 24^th^, 2020, more than 20 million cases and 800,000 deaths due to the disease were recorded globally (4).

In Greece, the first cases of COVID-19 were confirmed in late February, 2020 and within a period of one month, 1,212 people were diagnosed with the disease resulting in 46 deaths (5). The Greek Government took immediate action to minimize the spread of the virus by applying a series of public health measures, including the cancellation of local carnivals, closing of schools, universities, gyms, archaeological sites and eventually shopping malls, cafeterias, restaurants, bars and beauty parlors. From March 23^rd^ to May 4^th^, 2020, a mass lockdown was enforced, whereby citizens could leave their residences only under certain circumstances and after special notification (5).

From the onset of the COVID-19 pandemic, a large amount of information has been produced with respect to the biological features of the virus and the necessary measures of protection as well as the epidemiology of COVID-19. Human-to-human SARS-CoV-2 transmission is achieved primarily through droplets coming from infected person’s coughing or sneezing that can land directly on the mouth, nose, or eyes of a nearby person or on the surface of objects (6). Most common clinical symptoms of COVID-19 are loss of taste/smell, fever, cough, myalgia, fatigue and dyspnea (7). The rates of asymptomatic carriers are unknown with current estimations ranging from 40% to 45% and up to 62% of transmission may occur prior to the onset of symptoms (8,9). Evidence on the SARS-CoV-2 transmission by asymptomatic cases exists but relevant details still remain under discussion (10).

The need to control transmission necessitates the assessment of awareness, knowledge and adapted practices towards the new pandemic among the general population. Therefore, the primary objective of this study was to investigate the levels of knowledge and beliefs on the COVID-19 pandemic and the magnitude of trust upon Greek authorities and how these measures differed according to age, gender and time period among the participants of an ongoing Greek cohort study, the Epirus Health Study (EHS). We also explored possible factors associated with the population’s knowledge regarding SARS-CoV-2 transmission and seriousness of COVID-19 in a high-dimensional, agnostic manner to provide guidance for future public health policies tailored to certain population subgroups.

## METHODS

### Study population

The EHS (https://ehs.med.uoi.gr/) was initiated in June 2019 and is an ongoing population-based prospective cohort study, which aims to provide meaningful insight on the complex etiology of multifactorial chronic diseases and contribute to the improvement of the overall health state of the Greek population. The EHS cohort consists of permanent residents of the northwest region of Epirus in Greece, aged 25 to 70 years. Recruitment methods include advertisements to the local press and social media, promotions via study’s website, participation in events organized by local health agencies and invitations to Epirus residents and especially to the staff of companies of the private and public sector. Up to August 31^st^, 2020, a total of 667 subjects agreed to participate.

The study was approved by the Research Ethics Committee of the University of Ioannina and is conducted in accordance with the Declaration of Helsinki. All participants provide written informed consent prior to participation in the study.

### Data collection

#### Socio-demographic characteristics, general health status and lifestyle data

All participants were interviewed by two trained interviewers with the use of a standard questionnaire at recruitment. Information was collected regarding i) socio-demographic characteristics, such as age, gender, place of birth, marital status, educational level, employment status and income, ii) general health status, including symptoms of anxiety and depression, iii) personal and family medical history, and iv) lifestyle factors, including physical activity and sedentariness, smoking habits, alcohol consumption, cellphone use, sleep patterns, birth history, reproductive factors in women, cancer screening, medication and dietary supplement use, and dietary behaviors with a special focus on adherence to Mediterranean diet.

Symptoms of anxiety and depression were assessed with the two leading questions from the General Anxiety Disorder-7 (GAD-7) (11) and the Patient Health Questionnaire-9 (PHQ-9) (12), respectively. Adherence to Mediterranean diet was estimated by calculating the 14-point Mediterranean Diet Adherence Screener (MEDAS), which defines adherence as low (0-6 points) or high (7-14 points) (13). The participants’ sleep quality was assessed using the Pittsburgh Sleep Quality Index (PSQI) (14), which ranges from 0 to 21 and the higher the score, the poorer the sleep quality. Participants were classified as non-smokers, former smokers and current smokers according to their self-reported smoking habits. Finally, duration of recreational physical activity was assessed in days per week and minutes per day and then converted to metabolic equivalents of energy expenditure (MET). Each type of activity was assigned to a specific MET score, that is, 3.0 for walking, 6.0 and 9.0 for moderate-intensity and vigorous-intensity physical activity, respectively (15). The activity-specific MET value was then multiplied by the duration of activity in number of hours per week.

#### Anthropometric and clinical measurements

All participants attended a series of extensive clinical examinations at the baseline visit by two trained medical professionals. Weight and standing height were measured after removal of heavy clothing and shoes. Waist and hip circumference were measured at standing position after taking a deep breath at the thinnest spot of the waist and the widest spot of the hip, respectively. All anthropometric variables were measured using SECA equipment. Bioelectrical impedance analysis was performed using the Tanita MC-780MA machine. Pulse oxymetry was performed using the H-100B EDAN. Systolic and diastolic blood pressures were measured using the MicroLife A6 PC-AFIB PC monitor. Arterial stiffness was measured using the Mobil-O-Graph PWA New Generation 24h ABPM Classic monitor (16). In addition, cognitive function was assessed by three brief neuropsychological procedures, namely the Greek version of the Trail Making test (17,18), Logical Memory (19) and the Verbal Fluency test (20), and administered by two trained interviewers. Blood, urine and saliva samples were collected at recruitment after fasting for at least eight hours from all participants. Serum glucose (GLU), total cholesterol (TCHOL), low-(LDL) and high-density lipoprotein (HDL) cholesterol and triglycerides (TG) were measured at a commercial laboratory facility in Ioannina, Greece. Within- and between-lab variability was assessed on the same day on blind duplicate aliquots in a random sample of 11 and 7 participants, respectively. Duplicate aliquots were measured at the biochemistry laboratory of the University Hospital of Ioannina, Greece. The within-lab coefficients of variation (%CV) were 0.88%, 0.85%, 0.62%, 1.31% and 1.20% for GLU, TCHOL, LDL, HDL and TG, respectively, and 3.30%, 6.62%, 4.41%, 6.34% and 4.72% for the between-lab variability.

#### The COVID-19 sub-questionnaire

Due to the alarming spread of SARS-CoV-2 and the continuously increasing number of cases and subsequent deaths globally, a set of 24 COVID-19-related questions was integrated in the standard EHS questionnaire shortly after March 23^rd^, 2020, when lockdown measures were enforced in Greece, aiming to depict the level of awareness, knowledge and trust to the Greek authorities regarding the COVID-19 pandemic. Three questions investigated participants’ alertness regarding the pandemic and five questions assessed SARS-CoV-2 testing and COVID-19 clinical symptoms. Twelve questions evaluated the knowledge for the modes of SARS-CoV-2 transmission, severity of COVID-19 and participants’ capability of protecting themselves against SARS-CoV-2, and the last four questions examined participants’ beliefs towards the available information regarding the pandemic and trust in the Greek Government and health authorities for mitigating COVID-19. The COVID-19 sub-questionnaire was administered with face-to-face interviews, except for participants who joined the study before May 18^th^, 2020, for whom telephone-based interviews were conducted.

### Assessment of knowledge regarding the COVID-19 pandemic

A categorical variable was constructed to capture knowledge regarding the COVID-19 pandemic. Overall knowledge status was labeled as poor, moderate or good based on participants’ answers in the following items: modes of SARS-CoV-2 transmission, SARS-CoV-2 transmission by asymptomatic cases and COVID-19 severity. Poor knowledge was determined by answering “No” in at least one of the following three questions: i) “Is SARS-CoV-2 transmitted by droplets in the air?”, ii) “Is SARS-CoV-2 transmitted by contacting infected people?”, and iii) “Is SARS-CoV-2 transmitted by touching contaminated surfaces?”. When participants answered correctly to all three aforementioned questions, they were further asked whether an asymptomatic case could transmit the virus and their perceptive over COVID-19 severity. If the corresponding answers were “No” and “Many people survive, many people die”, “Some people survive, most people die” or “Almost everyone dies”, respectively, then the knowledge status was characterized as moderate. Finally, good knowledge was established if participants answered correctly to all aforementioned questions (**Additional File 1**).

### Statistical analysis

Participants’ baseline characteristics were summarized using means and standard deviations (SD) for continuous variables, and percentages for categorical variables. All questions included in the COVID-19 sub-questionnaire were summarized overall and by gender, age group (25-39, 40-49, 50-59, ≥60 years) and date of interview (March 30^th^ to May 3^rd^, 2020, and two post-lockdown time periods: May 4^th^ to June 31^st^, 2020 and July 1^st^ to August 31^st^, 2020). Pearson’s chi-square and Fisher’s exact tests were employed to detect differences between subgroups.

An agnostic exposure-wide association analysis was conducted using ordinal logistic regression models to quantify the associations between the level of participants’ knowledge towards the COVID-19 pandemic and 153 categorical and continuous explanatory variables arising from participants’ interviews and clinical examinations. Variables assessing verbal fluency were not considered eligible for our analysis due to high proportion of missing values. Dichotomous variables with prevalence of less than 10% and variables pertaining to a specific subgroup of participants (e.g. reproductive variables in women) were also excluded for statistical power reasons. Multiple comparisons were corrected using a false discovery rate (FDR) threshold of 5% based on the Benjamini–Hochberg approach (21). Continuous exposures and knowledge status results are presented as odds ratios (OR) per 1 SD increment. All models were adjusted for continuous age and sex. Statistical analyses were performed using STATA (version 14; StataCorp, College Station, TX, USA)

## RESULTS

### Participant characteristics

Of the 667 subjects enrolled in the EHS cohort up to August 31^st^, 2020, we excluded 105 (15.7%) participants who joined the study before March, 2020 and could not be reached via telephone to answer the COVID-19 sub-questionnaire. The distribution of baseline descriptive characteristics is shown in **Table 1**, and they were very similar between the EHS total and analytical sample. A total of 563 participants, 337 women and 226 men, constituted the analytical study sample, of them 170 women and 114 men completed the study before May 4^th^, 2020 when lockdown measures were partially lifted. The mean age of participants enrolled before or after May 4^th^, 2020 was 48.6 (SD=11.1) and 48.7 (SD=11.0) years, respectively. The majority (66.4%) of participants had university education, 31% were current smokers, 43% drank alcohol at least once per week, reported low levels of recreational physical activity and had a mean BMI of 26.6 kg/m^2^ (SD=4.7), and mean body fat percentage of 28.4% (SD=7.8) (**Table 1**).

**Table 1:** Descriptive characteristics of Epirus Health Study (EHS) participants

|  |  |  |
| --- | --- | --- |
|  | Total sample<br>(n=667) | Study sample<br>(n=563) |
| **Age** [years; mean (SD)] | 48.7 (11.3) | 48.7 (11.0) |
| **Gender** [n (%)] |  |  |
| Women | 399 (59.8) | 337 (59.9) |
| **Maximum level of education** [n (%)] |  |  |
| Junior high school | 67 (10.1) | 52 (9.3) |
| High school | 165 (24.8) | 136 (24.2) |
| University | 264 (39.6) | 228 (40.6) |
| Post-graduate studies | 170 (25.5) | 146 (26.0) |
| **Smoking status**<sup>1</sup> [n (%)] |  |  |
| Non-smokers | 297 (44.5) | 247 (43.9) |
| Former smokers | 163 (24.4) | 140 (24.9) |
| Current smokers | 207 (31.0) | 176 (31.3) |
| **Alcohol consumption** [n (%)] |  |  |
| Never | 58 (8.7) | 51 (9.1) |
| Less than once/month | 210 (31.5) | 177 (31.4) |
| 1-3 times/month | 110 (16.5) | 94 (16.7) |
| 1-2 times/week | 176 (26.4) | 152 (27.0) |
| At least 3 times/week | 113 (16.9) | 89 (15.8) |
| **Moderate-intensity recreational PA** [METs-hours/week, mean (SD)] | 5.3 (11.0) | 5.5 (11.5) |
| **Vigorous-intensity recreational PA** [METs-hours/week, mean (SD)] | 5.4 (15.4) | 4.9 (14.6) |
| **Mediterranean diet assessment score** [mean (SD)] | 7.2 (1.8) | 7.1 (1.8) |
| **Pittsburgh sleep quality index** [mean (SD)] | 5.1 (2.6) | 5.0 (2.4) |
| **Body mass index**<sup>2</sup> [kg/m<sup>2</sup>; mean (SD)] | 26.5 (4.7) | 26.6 (4.7) |
| **Body fat**<sup>2</sup> [%; mean (SD)] | 28.4 (7.8) | 28.4 (7.8) |
| **Systolic blood pressure**<sup>3</sup> [mmHg; mean (SD)] | 117.2 (15.1) | 117.5 (15.1) |
| **Diastolic blood pressure**<sup>3</sup> [mmHg; mean (SD)] | 75.0 (9.9) | 75.2 (9.9) |
| **Total cholesterol**<sup>4</sup> [mg/dL; mean (SD)] | 194.7 (34.1) | 194.7 (34.0) |
| **Low-density lipoprotein cholesterol**<sup>4</sup> [mg/dL; mean (SD)] | 126.4 (33.6) | 126.2 (33.5) |
| **High-density lipoprotein cholesterol**<sup>4</sup> [mg/dL; mean (SD)] | 54.3 (11.7) | 54.2 (11.8) |
| **Triglycerides**<sup>4</sup> [mg/dL; mean (SD)] | 94.6 (57.9) | 96.7 (59.7) |
| **Glucose**<sup>4</sup> [mg/dL; mean (SD)] | 85.5 (15.5) | 85.6 (15.9) |

*Abbreviations: METs, Metabolic equivalents of energy expenditure; PA, Physical activity; SD, Standard deviation*
<sup>1</sup>Variable indicating smoking status was constructed as follows: non-smokers; participants who were lifelong non-smokers or have tasted smoking products once or twice, former smokers; participants who have quitted smoking, current smokers; participants who smoked occasionally or were active smokers
<sup>2</sup>Parameters from lipometry tests conducted in the context of EHS
<sup>3</sup>Parameters measured using blood pressure monitors in the context of EHS
<sup>4</sup>Parameters from blood tests conducted in the context of EHS

### Awareness towards COVID-19 pandemic

In **Table 2**, COVID-19-related questions are presented in the total analytical sample and by gender, age group and period of interview. The vast majority of study participants (98.4%) were aware of the pandemic, although a significant difference was observed by gender as male participants showed slightly smaller awareness (96.9% vs 99.4%, p-value=0.034). Only 14 (2.5%) participants reported having a SARS-CoV-2 molecular test with none of them reporting a positive result. A total of 23 (4.5%) participants reported that they believed they had contracted the virus, although not tested, and 42 (7.5%) participants reported suffering from COVID-19-relevant symptoms during previous months, such as fever, cough, dyspnea and loss of taste/smell. The percentage of participants reporting belief of self-infection and symptoms of SARS-CoV-2 infection were higher for participants that joined the study during July and August, 2020 (15% and 21.7%, respectively) compared to previous months (p-value<0.001). Only two participants reported having a family member tested positive for SARS-CoV-2. Approximately 33% of participants considered themselves as absolutely capable of protecting themselves against SARS-CoV-2 with higher proportions observed in earlier times of interview (by time period: 43.6% vs. 23.4% vs. 19.2%, p-value<0.001), whereas 4.3% and 27.6% reported absolute and moderate certainty that they will not get infected, respectively, and the percentage of moderate certainty decreased with time (by time period: 33.3% vs. 27.4% vs. 14.2%, p-value<0.001) (**Table 2**).

**Table 2:**
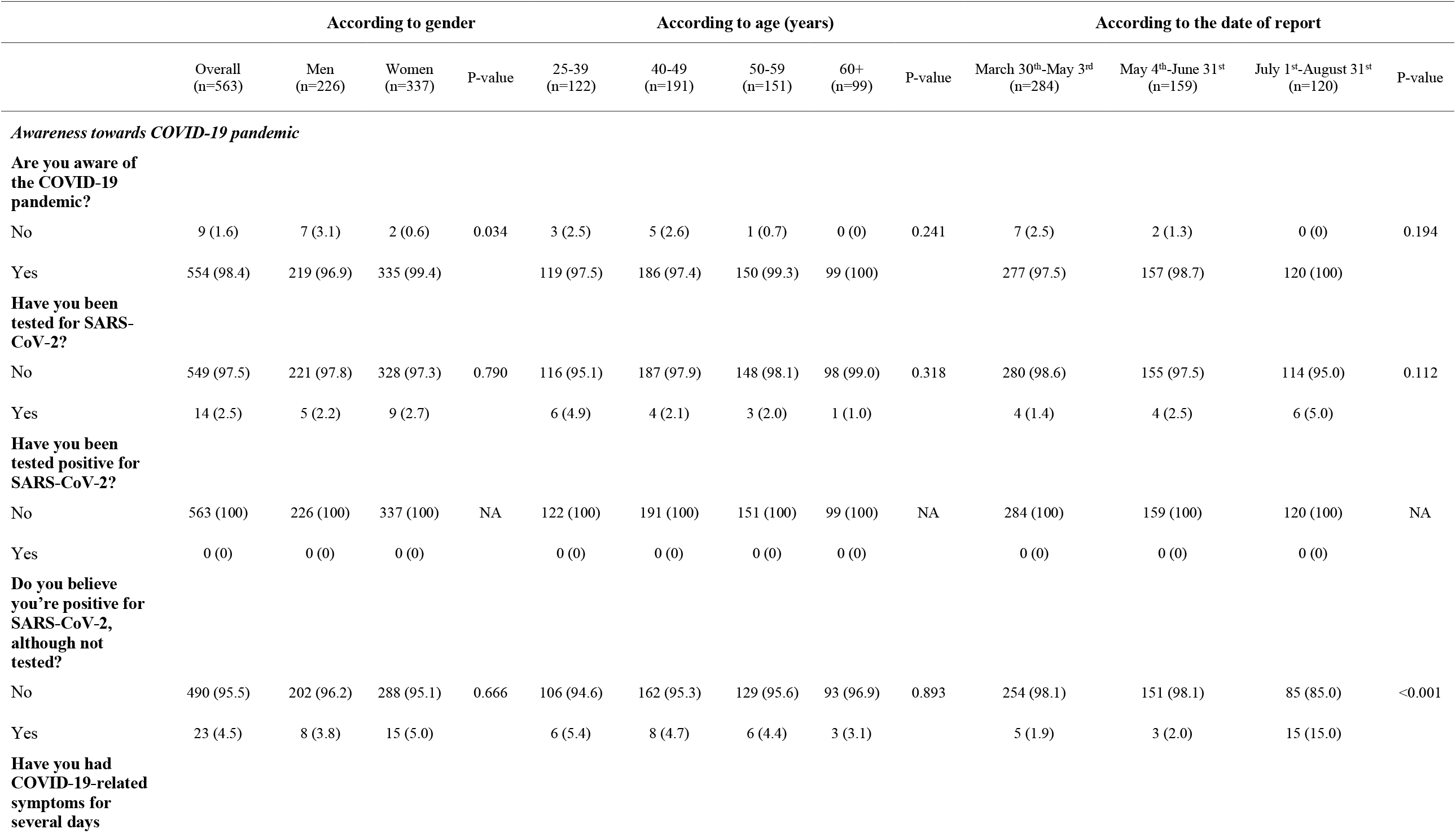
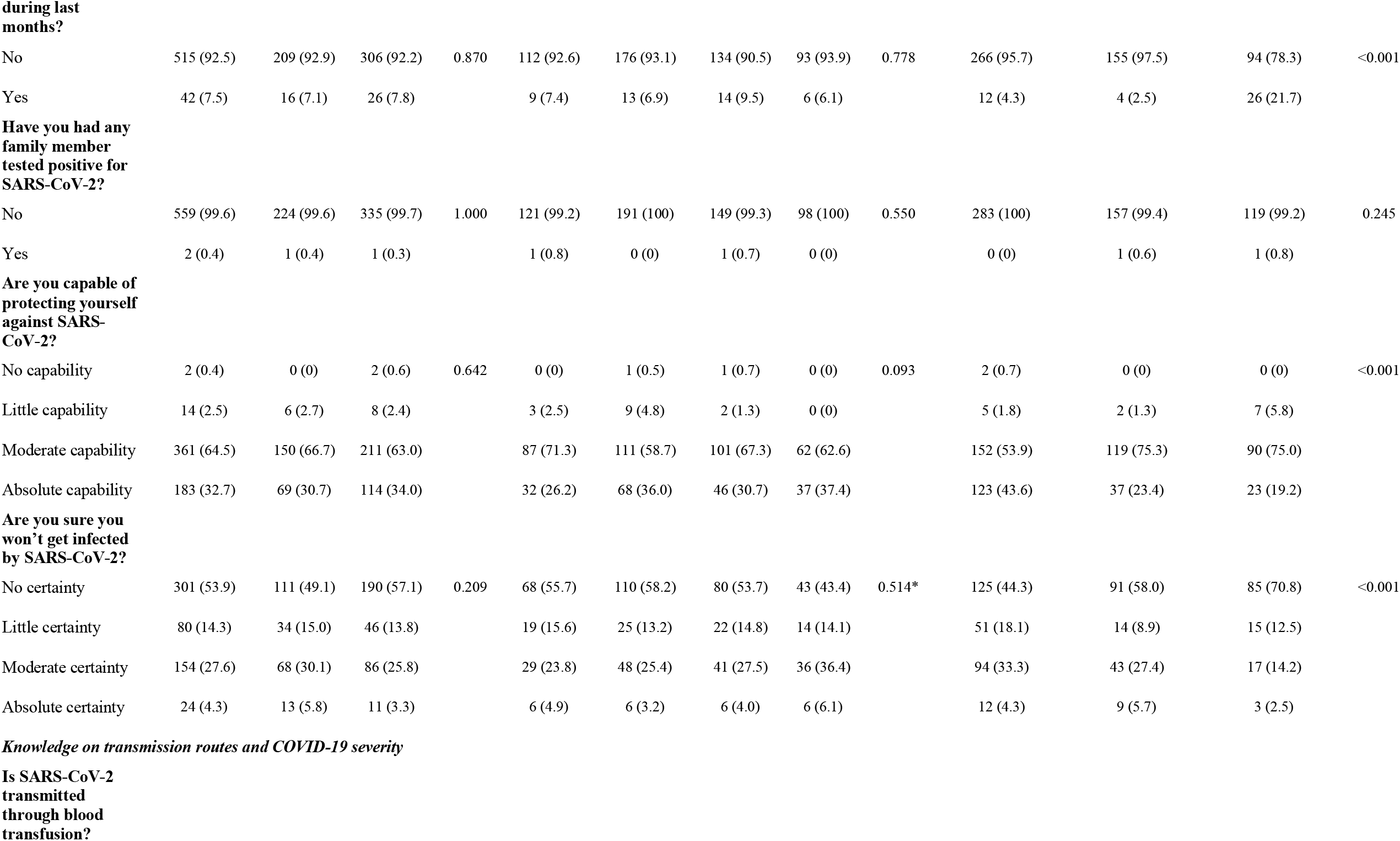
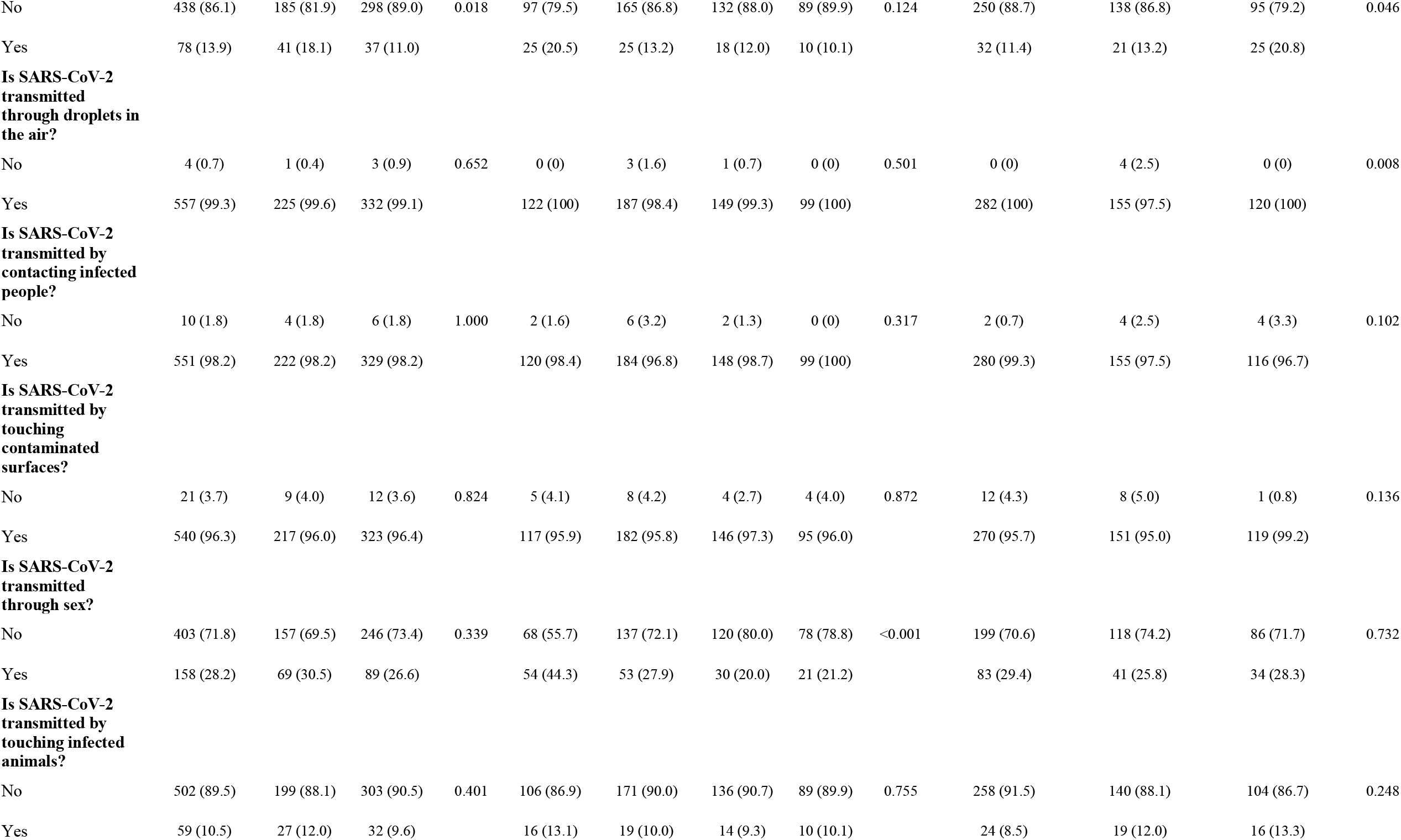
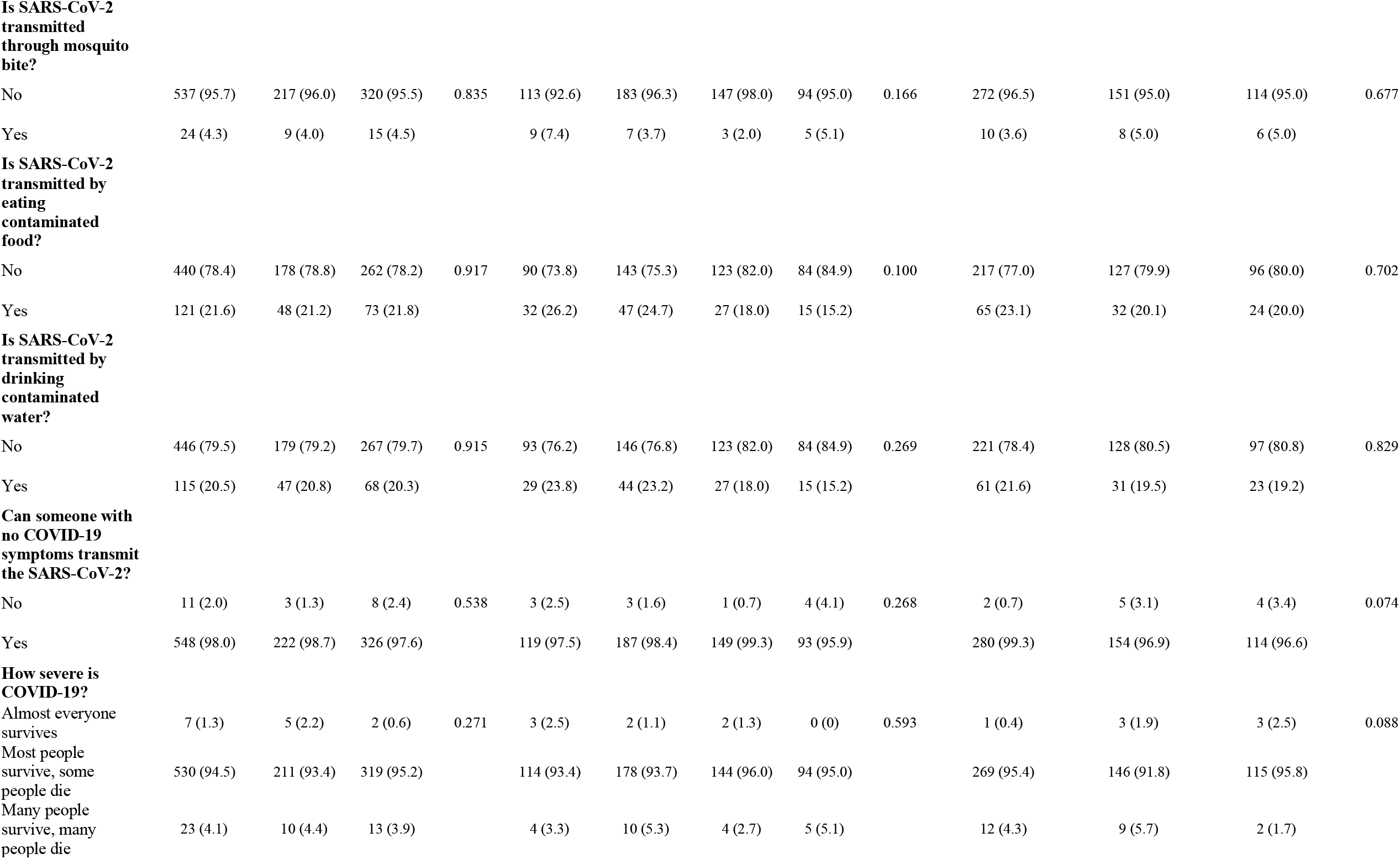
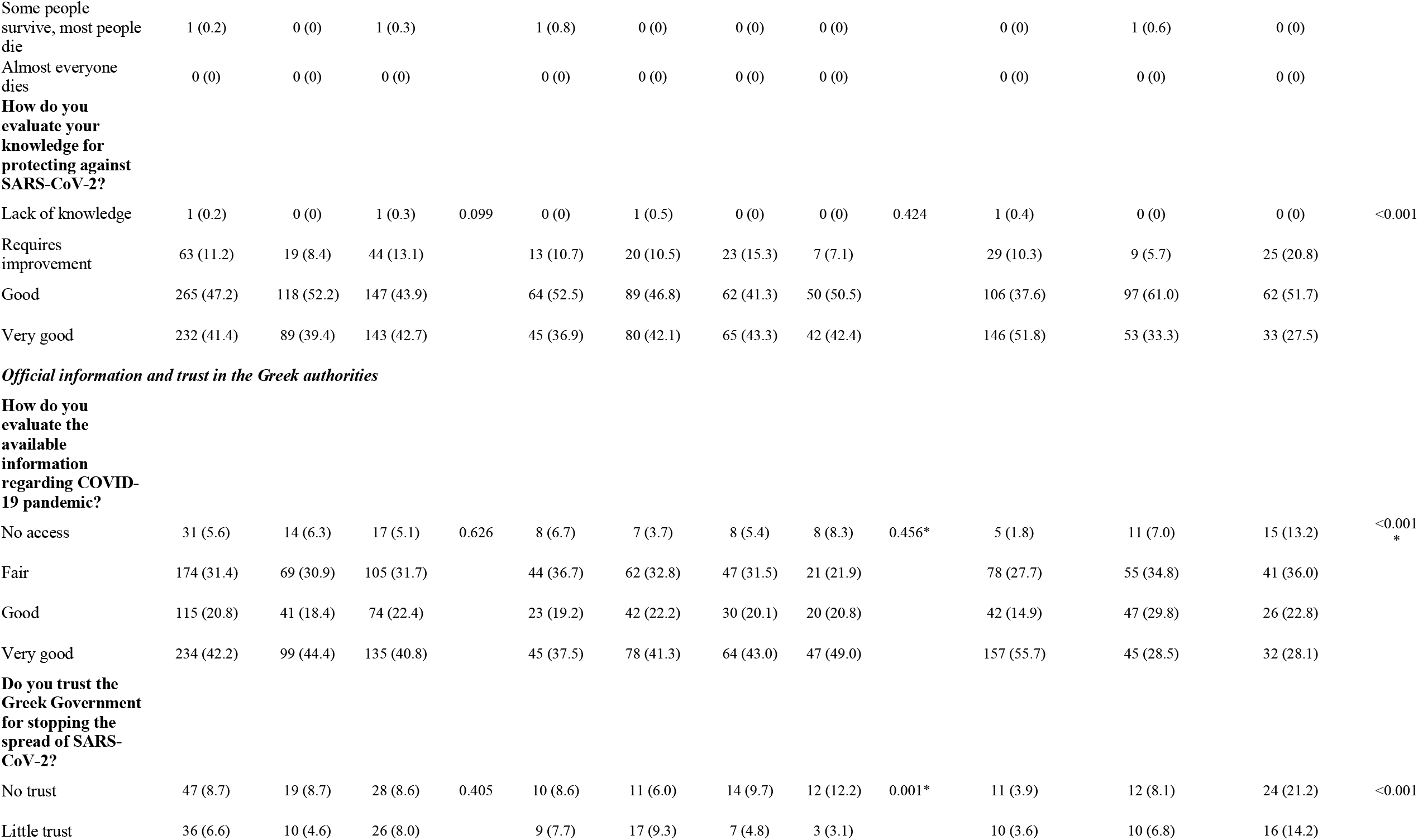
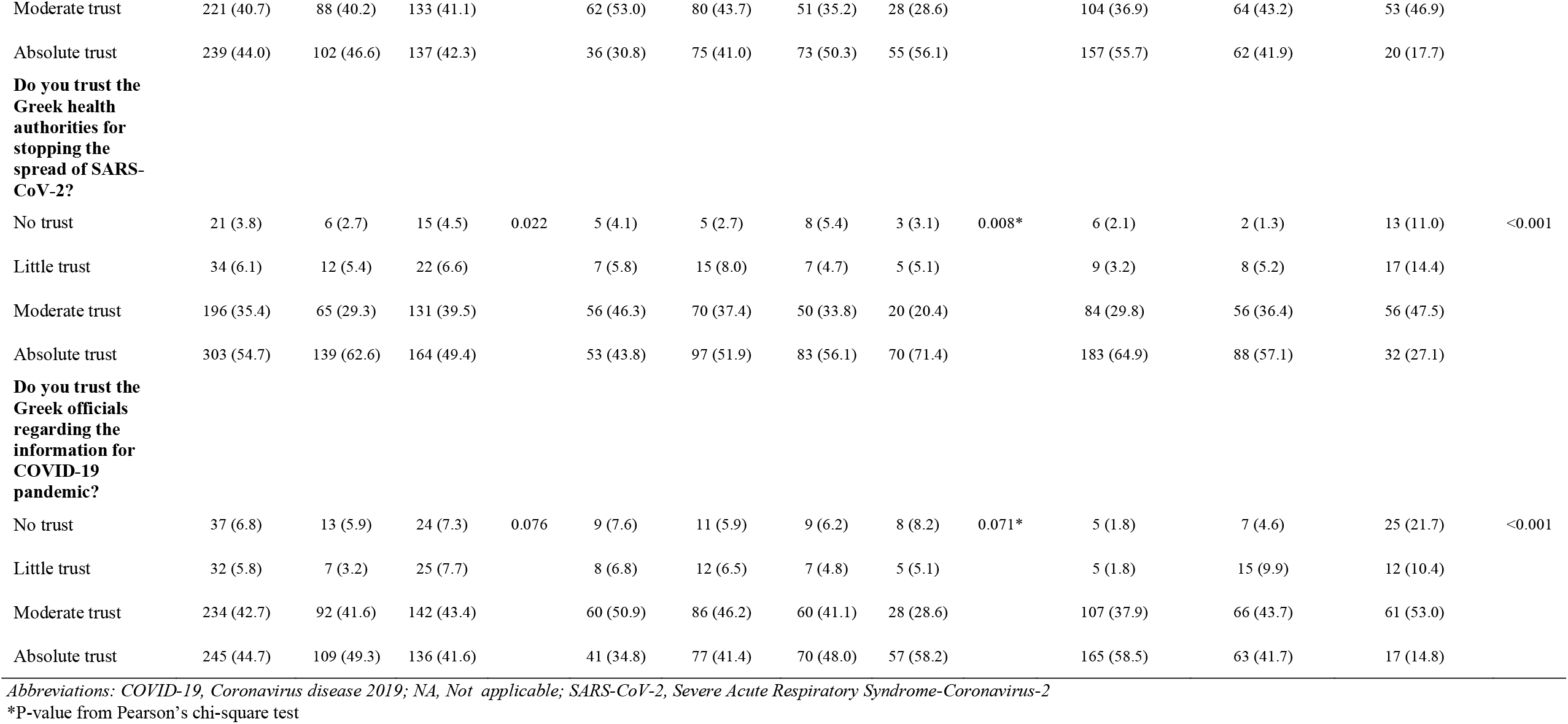
Frequency (N) and percentage (%) of Epirus Health Study (EHS) participants per COVID-19-related question overall and by gender, age group and date of report

### Knowledge on SARS-CoV-2 transmission routes and COVID-19 severity

More than 95% of participants reported that SARS-CoV-2 could be transmitted by droplets in the air, by contacting infected people or by touching contaminated surfaces (**Table 2**). Approximately 14%, 28% and 4% of participants reported that SARS-CoV-2 could be transmitted through blood transfusion, sexual intercourse or mosquito bite, respectively, and a larger proportion of participants younger than 50 years compared to older participants reported that SARS-CoV-2 can be transmitted through sexual intercourse (p-value<0.001). Almost all participants (98%) reported that SARS-CoV-2 could be transmitted by asymptomatic cases. Concerning COVID-19 severity, most participants (94.5%) reported that most people who contracted SARS-CoV-2 survive and some die (**Table 2**). When the questions regarding SARS-CoV-2 transmission and COVID-19 severity were aggregated together, approximately 86% of study participants demonstrated good level of knowledge without statistically significant differences between subgroups (**Table 3**). Yet still, almost 5% of study participants lacked basic knowledge regarding SARS-CoV-2 transmission routes. Similarly, a large proportion (88.3%) of participants declared that they had good or very good knowledge on how to protect themselves against SARS-CoV-2 with better knowledge associated with earlier interview periods (by time period: 89.4% vs. 94.3% vs. 79.2%, p-value<0.001) (**Table 2**).

**Table 3:** Frequency (N) and percentage (%) of Epirus Health Study (EHS) participants according to the level of knowledge for the COVID-19 pandemic overall and by gender, age group and date of report

|  | According to gender |  |  |  | According to age (years) |  |  |  |  | According to the date of report |  |  |  |
| --- | --- | --- | --- | --- | --- | --- | --- | --- | --- | --- | --- | --- | --- |
|  | Overall<br>(n=563) | Men<br>(n=226) | Women<br>(n=337) | P-value<sup>†</sup> | 25-39<br>(n=122) | 40-49<br>(n=191) | 50-59<br>(n=151) | 60+<br>(n=99) | P-value<sup>†</sup> | March 30<sup>th</sup>-May 3<sup>rd</sup><br>(n=284) | May 4<sup>th</sup>-June 31<sup>st</sup><br>(n=159) | July 1<sup>st</sup>-August 31<sup>st</sup><br>(n=120) | P-value<sup>†</sup> |
| Poor knowledge | 25 (4.5) | 9 (4.0) | 16 (4.8) | 0.860 | 5 (4.1) | 11 (5.8) | 6 (4.0) | 3 (3.0) | 0.877 | 12 (4.3) | 9 (5.7) | 4 (3.3) | 0.140 |
| Moderate knowledge | 56 (10.0) | 24 (10.6) | 32 (9.6) |  | 14 (11.5) | 17 (9.0) | 13 (8.7) | 12 (12.1) |  | 20 (7.1) | 22 (13.8) | 14 (11.7) |  |
| Good knowledge | 480 (85.6) | 193 (85.4) | 287 (85.7) |  | 103 (84.4) | 162 (85.3) | 131 (87.3) | 84 (84.9) |  | 250 (88.7) | 128 (80.5) | 102 (85.0) |  |

Abbreviation: COVID-19, Coronavirus disease 2019
<sup>†</sup>P-value from Fisher’s exact test

### Exposure-wide association analysis on knowledge status towards COVID-19 pandemic

Only 12 (7.8%) explanatory variables were found significantly associated with the level of participants’ knowledge towards the COVID-19 pandemic at the 0.05 level (see **Figure 1** and **Additional File 2**). Odds of having a lower knowledge status were associated with higher extracellular water-to-total body water ratio, increased incapability to fall asleep within 30 minutes during last month, frequent talking on cellphone away from the ear during last three months, lower self-reported health status, higher heart rate, frequent pain at night during last month, having family members diagnosed with acute depression or osteoporosis/hip fracture and limited interest for personal or professional activities during last two weeks. Higher pulse volume, increased trust in national health authorities to limit spread of the virus and increased paid hours of work per week were associated with higher level of knowledge. However, none of these associations were significant after correcting for multiple comparisons (**Figure 1** and **Additional File 2**).

**Figure 1:**
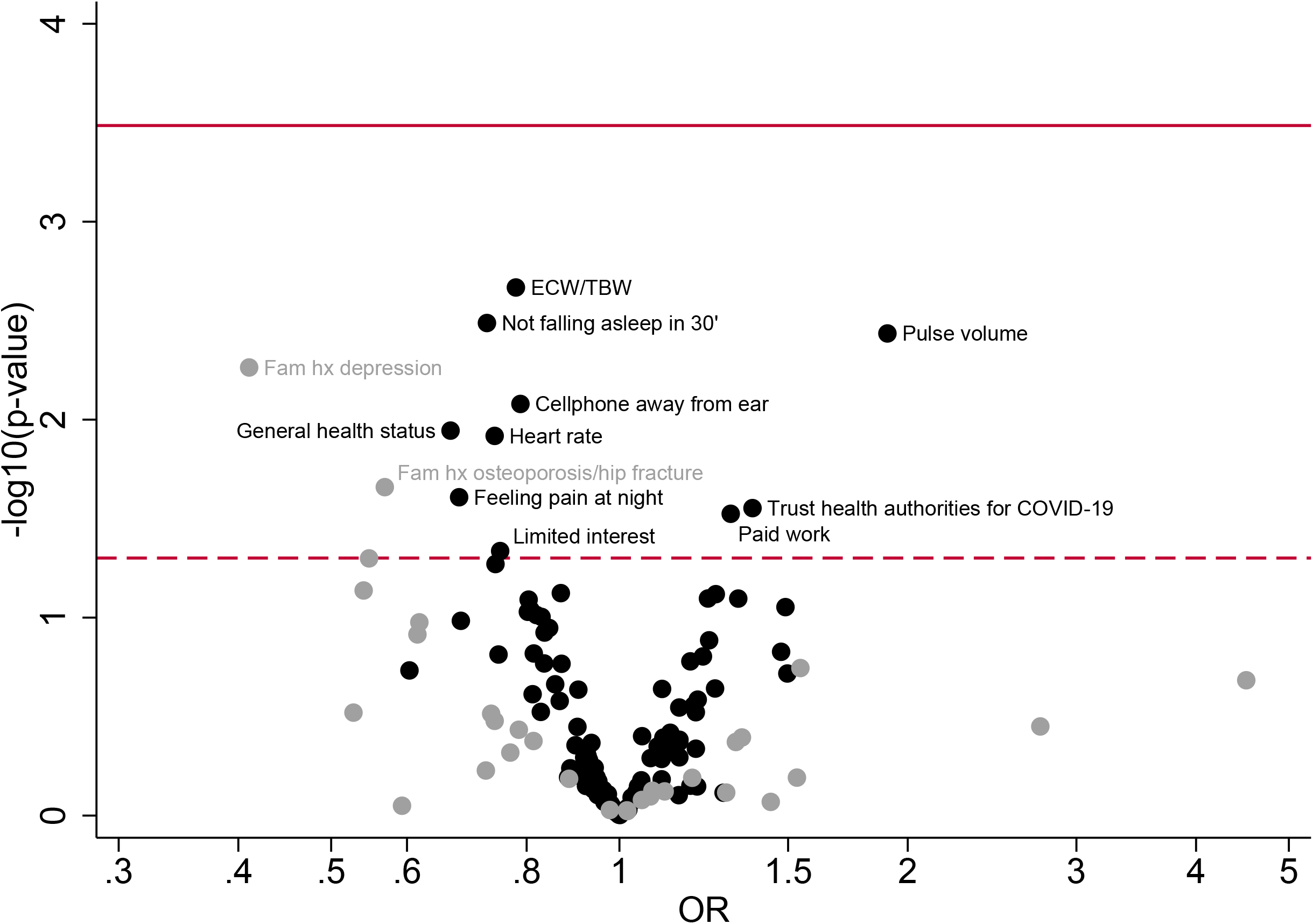
Volcano plot showing results from the exposure-wide association study regarding the association between 153 clinical parameters, medical characteristics, demographic and lifestyle factors and knowledge regarding the SARS-CoV-2 transmission routes and COVID-19 severity in the Epirus Health Study cohort. The *Y*-axis shows the p-values in −log10 scale from the ordinal logistic regression models for each factor. The *X*-axis shows the estimated odds ratios for each factor. All models were adjusted for continuous age and gender. The black points indicate the continuous/ordinal exposures and the grey points indicate the categorical exposures. The dashed horizontal line represents the nominal level of statistical significance (0.05) and the solid horizontal line represents the FDR-corrected level of statistical significance (0.0003268). *Abbreviation:* COVID-19; Coronavirus disease 2019, ECW/TBW; Extracellular water-to-total body water ratio. OR; Odds ratio

### Trust in the Greek authorities for minimizing the spread of SARS-CoV-2

Publicly available information regarding COVID-19 pandemic was characterized as good (20.8%) and very good (42.2%) by more than half of the study sample (**Table 2**), but the responses differed by time period with more participants reporting better quality of information when interviewed before compared to after May 4^th^, 2020 (by time period: 70.6% vs. 58.3% vs. 50.9%, p-value<0.001). The percentage of study participants who reported absolute and moderate trust in the Greek Government for mitigating the spread of the virus was 44% and 40.7%, respectively; these percentages increased with higher age of participants (p-value=0.001) and were higher with earlier time of interview (absolute trust by time period: 55.7% vs. 41.9% vs. 17.7%, p-value<0.001). A similar pattern was observed with respect to trusting the Greek health authorities for mitigating the spread of the virus and the official Greek sources of information regarding COVID-19 pandemic (**Table 2**). When the weekly moving average of COVID-19 cases and deaths in Greece were plotted versus the weekly moving average percentage of EHS participants who reported absolute or moderate trust in the Greek authorities, participants’ trust seemed to decrease gradually with time without following the pattern of COVID-19 cases in Greece (**Figure 2**).

**Figure 2:**
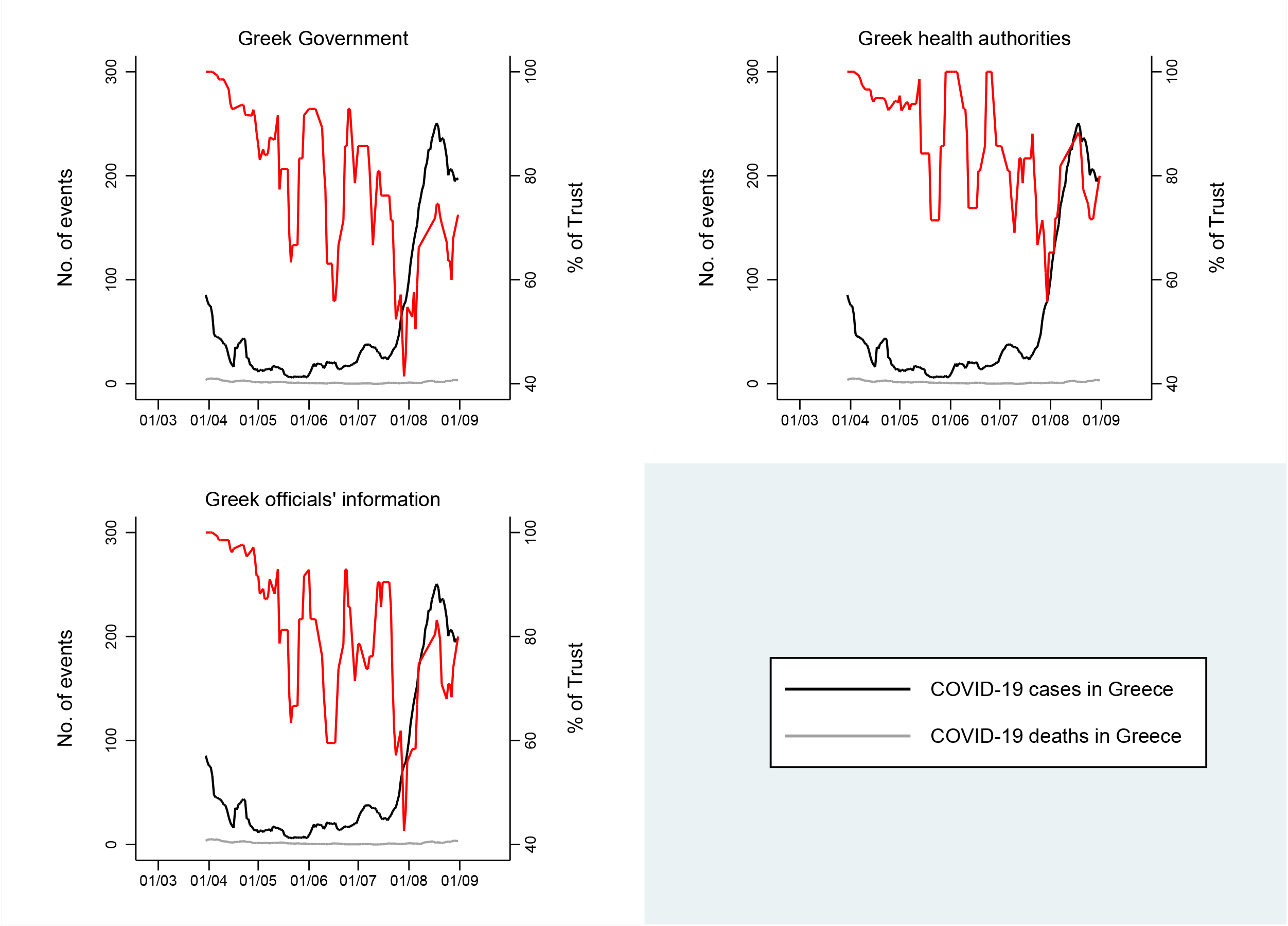
Weekly moving average of COVID-19 confirmed cases and deaths in Greece along with the weekly moving average percentage of participants who reported absolute or moderate trust in the Greek Government (panel A), Greek health authorities (panel B) and Greek officials’ information (panel C) in the Epirus Health Study cohort. The left *Y*-axis represents absolute frequencies and the right *Y*-axis represents percentages. Data on daily COVID-19 cases and deaths in Greece was obtained from the National Public Health Organization. *Abbreviation: COVID-19; Coronavirus disease 2019*

## DISCUSSION

This study evaluated the degree of knowledge about the COVID-19 pandemic and the trust upon the official authorities from March 30^th^ to August 31^st^, 2020 in 563 participants of an ongoing cohort study in Greece. The participants demonstrated high levels of knowledge regarding SARS-CoV-2 transmission and COVID-19 severity without statistically significant differences by age, gender and time of interview. High levels of trust in the Greek authorities for providing information and mitigating the spread of SARS-CoV-2 were also observed, which were stronger in older participants and those that joined the study closer to the start of the pandemic. We also examined in an agnostic fashion 153 potential correlates of the level of knowledge about the pandemic, but no associations survived the multiple testing correction.

Participants were well-informed on the modes of SARS-CoV-2 transmission via respiratory droplets, contacting infected people or touching contaminated surfaces. The vast majority of them also understood risk of infection caused by asymptomatic cases and the severity of COVID-19. Only a small percentage of participants reported possibility of transmission through blood transfusion and mosquito bites, but 28% of them reported that the virus could be transmitted by sexual intercourse. Even though medical professionals understand sexual transmission as a separate entity, the general public may still consider the sexual intercourse related risk of SARS-CoV-2 infection as possible due to the presumed risk of infection through respiratory droplets that takes place during sexual intercourse. Literature evidence on the knowledge and attitudes of the Greek population towards the COVID-19 pandemic is limited. In a sample of 461 health care professionals working in five public hospitals in the central region of Thessaly, Greece recruited in February, 2020, 96% recognized that droplets is the main mode of SARS-CoV-2 transmission, whereas 20% and 53% considered sex and food consumption, respectively, as possible modes of transmission (22). Our findings are compatible with several studies conducted in various countries worldwide, in which participants showed good knowledge for the transmission routes and the possible risk of infection imposed by asymptomatic people (23–30). However, most studies recruited targeted populations, including health care personnel, students and hospital visitors; thus, the generalization of their findings to the general public was questionable.

When we further investigated the association between participants’ level of knowledge and a variety of clinical parameters, medical history, demographic and lifestyle factors, positive associations emerged at the nominal statistical significance level between participants’ level of knowledge and trust in the Greek health authorities for managing the spread of the virus and the hours of paid work per week. Poorer health status, certain sleep disorder symptoms, higher heart rate and limited interest for participating in social and professional activities were inversely associated with higher knowledge status that could be explained by poorer ability to process relevant medical information. After adjustment for multiple testing, none of these factors remained significant (smallest FDR-adjusted p-value= 0.19). The relatively small sample size and the lack of large variability across participants’ level of knowledge may partially explain our null results.

The majority of participants expressed high levels of trust in the Greek authorities for providing information and mitigating the spread of SARS-CoV-2. These results might be attributed to the immediacy of the Greek Government on enacting strict measures shortly after the registration of the first cases of COVID-19 in Greece. Previous studies have reported mixed results for participants’ trust in politicians and governmental authorities during the pandemic. Online surveys contacted in the USA and African countries suggested that the proportion of participants who showed trust in the corresponding Governments ranged from 10.2% to 38.6% (31–33). On the other hand, samples of Malaysian, Australian and Israeli residents considered governmental authorities as credible and capable to handle the COVID-19 health crisis (28,34,35). These contradictory findings highlight the magnitude in which the different strategies followed worldwide for the management of the pandemic and the resulting pandemic severity affect public standpoint.

Stratified analyses by time period of interview showed that the high levels of trust in the Greek authorities to mitigate spread of the virus waned with time, as the percentage of absolute and moderate trust in the Greek Government and health authorities decreased from approximately 92% and 94% in March/April, 2020 to 61% and 73% in July/August, 2020, respectively. Visual inspection of the trends did not provide any insights on a potential correlation between the weaning trust and the patterns of the number of confirmed COVID-19 cases or the subsequent deaths in Greece. Potential explanations for the gradual decline in participants’ trust could be the negative economic and perceived health impact of lockdown measures on post-lockdown participants’ beliefs and trust upon officials, and the resurgence of the epidemic in August, 2020 in Greece.

This study is one of the first population-based studies conducted in Greece that not only attempted to shed light on participants’ perceptions surrounding the COVID-19 pandemic and trust upon authorities but also evaluated trends by age, gender and time of assessment that can provide guidance for future public health policies tailored to certain population subgroups. Another strength of the study includes that the COVID-19-related data were collected as part of an ongoing prospective and heavily phenotyped epidemiological study, which provided the ability to systematically examine a number of potential factors and correlates in relation to participants’ level of knowledge for the pandemic. Several limitations should also be considered in interpreting our findings. The sample size was relatively small, but several of our findings have been previously observed in larger studies. The study sample constituted only by residents of the region of Epirus and had a higher participation of women and individuals with university education; thus, findings might not be generalizable to the general public. As a validated questionnaire to assess knowledge on the COVID-19 pandemic is not yet available, we constructed an aggregated variable to assess knowledge of SARS-CoV-2 transmission and COVID-19 severity that showed relatively poor variation, as most individuals reported good levels of knowledge.

## CONCLUSIONS

In the current study, participants showed an advanced level of knowledge of SARS-CoV-2 transmission and COVID-19 severity, and high levels of trust in the Greek authorities for providing information and mitigating the spread of SARS-CoV-2. The level of trust in the authorities weakened with time, which might highlight the importance of following evidence-based decision-making processes in managing the pandemic, and developing efficient collaborations between public health officials and Governments to sustain continued, accurate and higher effort information campaigns to increase public awareness.

## Data Availability

Data are available upon reasonable request to the corresponding author.

## LIST OF ABBREVIATIONS

CI: Confidence interval
%CV: Coefficient of variation
COVID-19: Coronavirus disease 2019
ECW-to-TBW: Extracellular water-to-total body water ratio
EHS: Epirus Health Study
FDR: False discovery rate
GAD-7: General Anxiety Disorder-7
GLU: Glucose
HDL: High-density lipoprotein cholesterol
LDL: Low-density lipoprotein cholesterol
MEDAS: Mediterranean Diet Adherence Screener
MERS-CoV: Middle East Respiratory Syndrome-Coronavirus
MET: Metabolic Equivalent of Energy Expenditure
NA: Not applicable
OR: Odds ratio
PHQ-9: Patient Health Questionnaire-9
PSQI: Pittsburgh Sleep Quality Index
SARS-CoV-2: Severe Acute Respiratory Syndrome-Coronavirus-2
SARS-CoV: Severe Acute Respiratory Syndrome-Coronavirus
SD: Standard deviation
TCHOL: Total cholesterol
TG: Triglycerides
USA: United States of America
WHO: World Health Organization

## DECLARATIONS

### Ethics approval and consent to participate

Written informed consent was obtained from all participants. The study protocol was approved by the Research Ethics Committee of the University of Ioannina, Ioannina, Greece.

### Consent for publication

Not applicable.

### Availability of data and materials

Data are available upon reasonable request to the corresponding author.

### Funding

This study was funded by the Operational Programme Epirus 2014-2020 of the Prefecture of Epirus

(HΠ1AB-0028180).

## Authors’ contributions

KKT, KV, EN, ECR, EE, IT, MTD, DES, VTT designed the Epirus Health Study. AK, FK, GM, EB, and KKT designed the current investigation; AK performed the statistical analysis, interpreted the results, drafted the first version of the manuscript and all authors revised the manuscript, discussed the results, and commented on the drafts; KKT had the primary responsibility for final content. All authors read and approved the final manuscript.

## Acknowledgements

Not applicable.

