## Supplementary figures and images for "Awareness, knowledge and trust in the Greek authorities towards COVID-19 pandemic: results from the Epirus Health Study cohort"

### Additional File 1

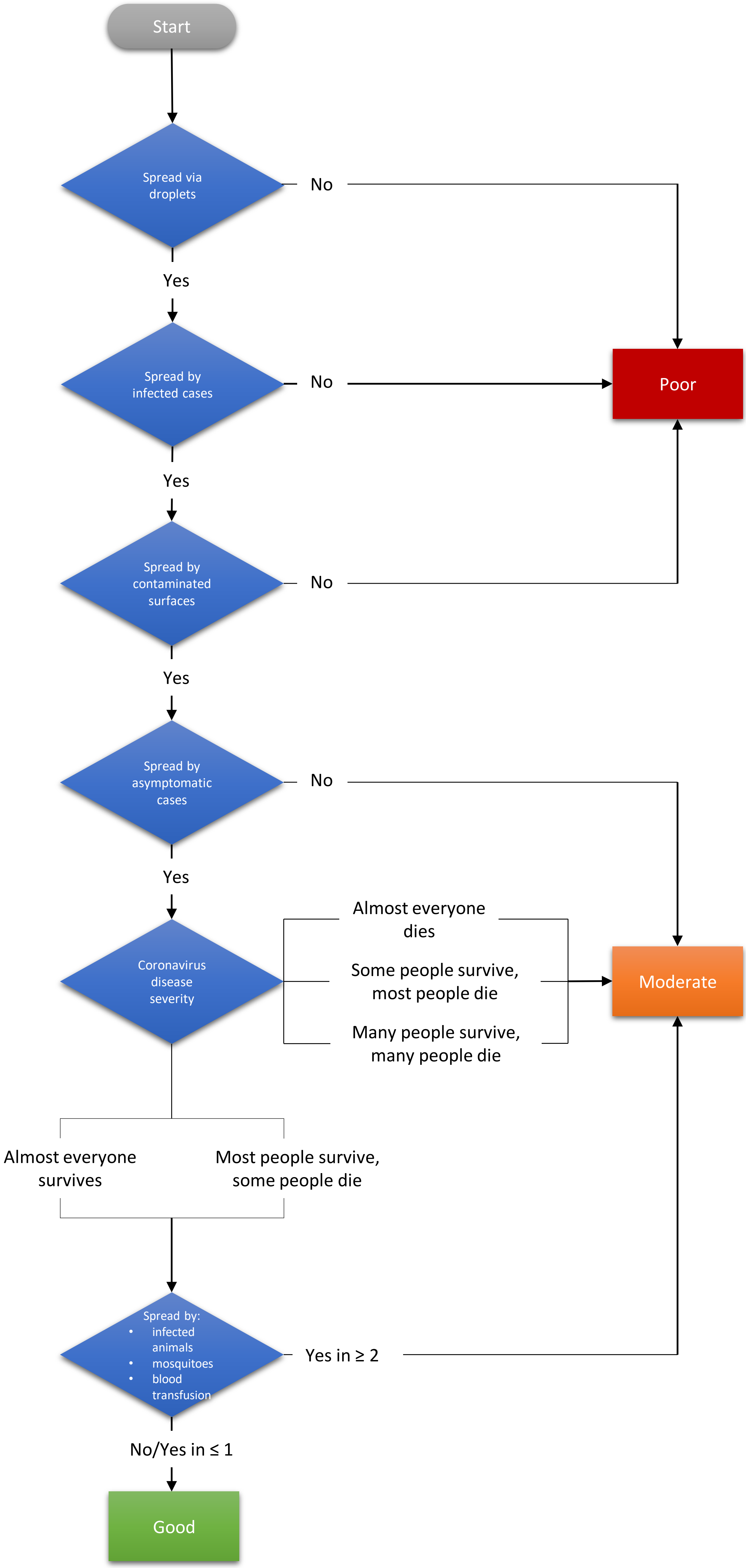
